# Seasonality of Common Human Coronaviruses in the United States, 2014-2021

**DOI:** 10.1101/2022.03.22.22272640

**Authors:** Melisa M. Shah, Amber Winn, Rebecca M. Dahl, Krista L. Kniss, Benjamin J. Silk, Marie E. Killerby

## Abstract

The four common human coronaviruses (HCoVs), including two alpha (HCoV-NL63 and HCoV-229E) and two beta (HCoV-HKU1 and HCoV-OC43) types, generally cause mild, upper respiratory illness. HCoVs are known to have seasonal patterns and variation in predominant types each year, but defined measures of seasonality are needed. We defined seasonality of HCoVs during July 2014 to November 2021 in the United States using a retrospective method applied to National Respiratory and Enteric Virus Surveillance System (NREVSS) data. In the six HCoV seasons prior to 2020-2021, onsets ranged from October to November, peaks from January to February, and offsets from April to June; most (>93%) HCoV detections occurred within the defined seasonal onsets and offsets. The 2020-2021 HCoV season onset was delayed by 11 weeks compared to prior seasons, likely due to COVID-19 mitigation efforts. Better defining HCoV seasonality can inform clinical preparedness and the expected patterns of emerging HCoVs.

**Article Summary Line:** The typical common HCoV season in the United States starts between October and November, peaks towards the end of January, and ends between April and June, but the 2020-2021 season was markedly delayed compared to prior seasons.

## Background

The four common human coronaviruses (HCoVs), including two alpha (HCoV-NL63 and HCoV-229E) and two beta (HCoV-HKU1 and HCoV-OC43) types, generally cause mild, upper respiratory illness. The HCoVs are endemic in humans, as evidenced by sustained, widespread, continuous transmission, unlike SARS-CoV and Middle East respiratory syndrome coronavirus (MERS-CoV), which are betacoronaviruses first detected in 2002 and 2012, respectively. HCoVs are known to circulate annually in the United States with a seasonal pattern, generally peaking between December and March with variation in predominant types each year (1). An additional coronavirus, SARS-CoV-2, emerged in the human population in late 2019 and has become widespread.

While HCoVs are known to have seasonal patterns, parameters of expected seasonality have not been defined. Given mitigation efforts and behavior changes due to the COVID-19 pandemic, national patterns of respiratory viruses including influenza differed during the 2020-2021 season compared to previous seasons (2). Understanding changes to seasonal patterns in HCoV circulation is important for clinical and public health preparedness and may provide insight into transmission patterns for novel HCoVs. Here we describe laboratory results of four common HCoVs in the United States from July 2014 to November 2021 using data collected by the National Respiratory and Enteric Virus Surveillance System (NREVSS).

## Methods

We analyzed circulation of HCoVs (hereafter defined as HCoV-NL63, HCoV-229E, HCoV-HKU1, and HCoV-OC43, and excluding SARS-CoV-2) using data from the National Respiratory and Enteric Viruses Surveillance System (NREVSS), a passive surveillance system established by the Centers for Disease Control and Prevention (CDC) in the 1980s. NREVSS collects respiratory virus testing results from laboratories across the United States (www.cdc.gov/surveillance/nrevss/labs/map.html). Clinical, public health, and commercial laboratories submit weekly aggregated numbers of tests performed and numbers of positive reverse transcriptase polymerase chain reaction (RT-PCR) detections for the four common HCoV types.

We included reports of specimens tested for HCoVs from the week ending July 5, 2014 to the week ending November 6, 2021. We excluded HCoV results without typing and data from laboratories that did not report any positive HCoV test results during the study period. We compiled total HCoV testing and positive detections by type, season, and U.S. Census region. We used a subset of data submitted through the Public Health Laboratory Interoperability Project (PHLIP) with specimens tested for all four HCoV types collected between June 29, 2014 and November 29, 2021 to characterize detections by age and sex. We also examined codetections with other respiratory viruses, including parainfluenza viruses (PIV), respiratory syncytial virus (RSV), human metapneumovirus (HMPV), human adenovirus (HAdV), rhinovirus/enterovirus (RV/EV), and influenza A and B in the PHLIP subset. We excluded specimens with pan-positive results for codetections in the PHLIP subset.

We evaluated the onset and offset of seasons between MMWR week 31 (early August) through MMWR week 30 of the following year (https://ndc.services.cdc.gov/wp-content/uploads/MMWR_Week_overview.pdf) using a method previously validated for RSV detections from NREVSS. This method (Retrospective Slope 10 method) is characterized by a centered, 5-week moving average of weekly detections with each seasonal peak normalized to 1,000 detections (3). The absolute difference between normalized detections for each week and the week prior was determined. The season onset was defined as the second consecutive week with an absolute difference of ≥10 normalized detections. The offset was defined as the last of two consecutive weeks when the number of normalized detections was greater than the number of normalized detections during the onset week. We determined seasonal characteristics nationally, including season onset, peak, and offset as well as season duration and the percentage of annual detections that occurred within the season.

Analysis was performed using RStudio version 1.4.1106. This activity was reviewed by CDC and conducted consistent with applicable federal law and CDC policy (45 C.F.R. part 46.102(l)(2), 21 C.F.R. part 56; 42 U.S.C. Sect. 241(d); 5 U.S.C. Sect. 552a; 44 U.S.C. Sect. 3501 et seq.)

## Results

Any HCoV type was detected in 104,911 (3.6%) of 2,878,479 specimens with results submitted to NREVSS during the weeks ending July 5, 2014 through November 6, 2021. Among these 104,911 specimens, 40.1% were HCoV-OC43-positive, 27.8% were HCoV-NL63-positive, 19.9% were HCoV-HKU1-positive, and 12.2% were HCoV-229E-positive. Testing volumes from NREVSS during March 2020, the onset of the COVID-19 pandemic, were higher than any other season (Figure 1A). The predominant common HCoV type fluctuated by surveillance year (Figure 1B).

**Figure 1:**
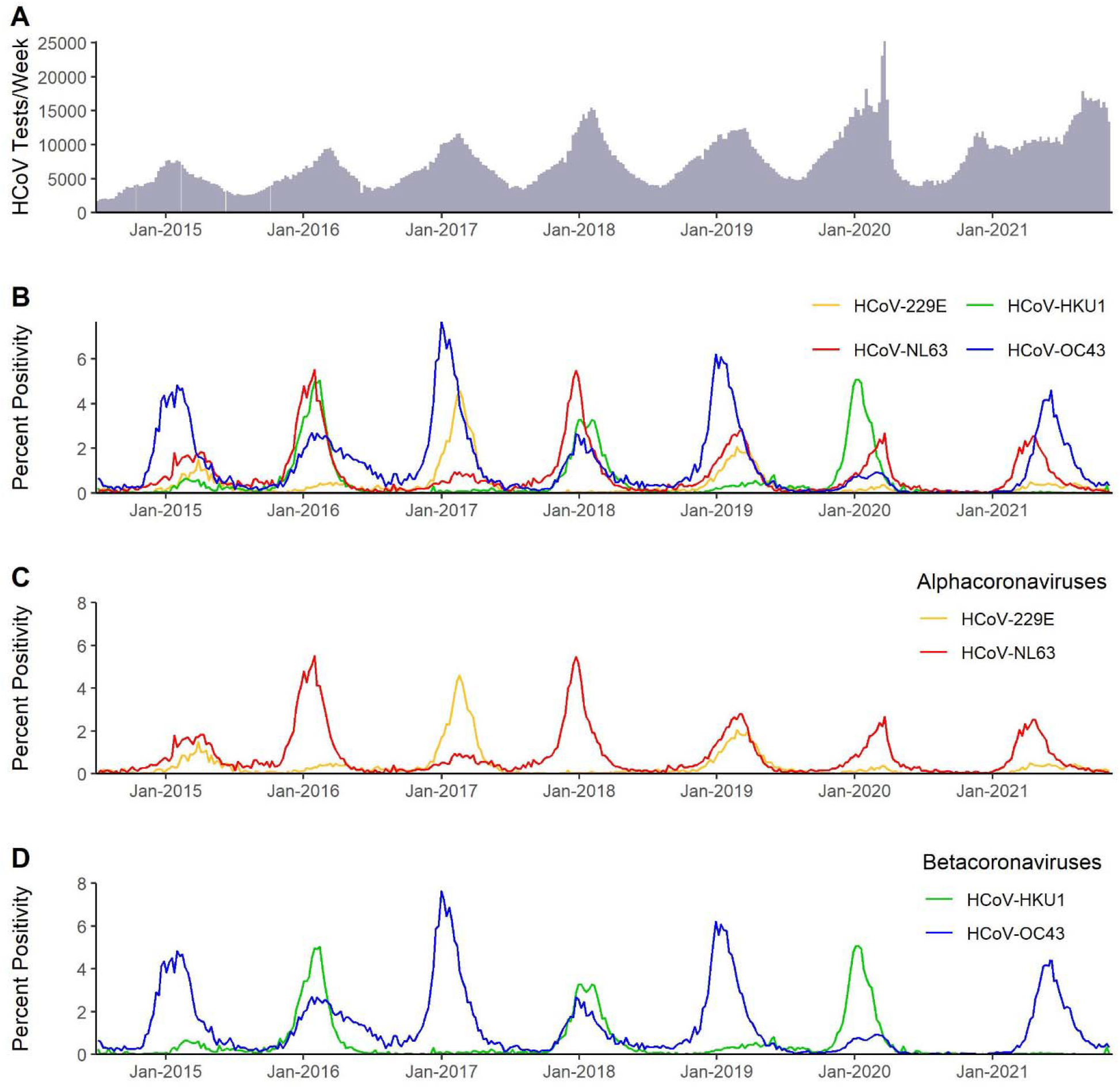
Total tests and percent positivity of four common human coronaviruses (HCoVs) in the United States from weekly aggregated data submitted to the National Respiratory and Enteric Virus Surveillance System (NREVSS), July 2014-November 2021. Panel A represents total specimens tested for all four HCoV types. Panel B shows percent positivity of the four HCoV types by week, and Panels C and D show percent positivity of the common alphacoronaviruses and common betacoronaviruses respectively.

In the six HCoV seasons before the COVID-19 pandemic (i.e., excluding the 2020-2021 season), seasonal onsets occurred between October and November, peaks occurred between January and February, and seasonal offsets occurred between April and June (Figure 2, Table 1). Specifically, the seasonal onset, peak, and offset occurred on average in MMWR week 44 (range: weeks 42 to 45), week 4 (range: weeks 1 to 6), and week 19 (range: weeks 17 to 25), respectively (Figure 3, Table 1); 93.2% of all HCoV detections occurred between the onset and offset. The mean duration of the 6 seasons prior to 2020-2021 was 25 weeks. The 2020-2021 common HCoV season onset was delayed by 11 weeks compared to prior seasons (Figure 3, Table 1). The number of days between onset and peak for the 2020-2021 season was longer than the mean observed for the prior 6 seasons (119 days versus 88 days, respectively). By November of 2021, normalized values had not reached the requirement for offset for the 2020-2021 season.

**Figure 2:**
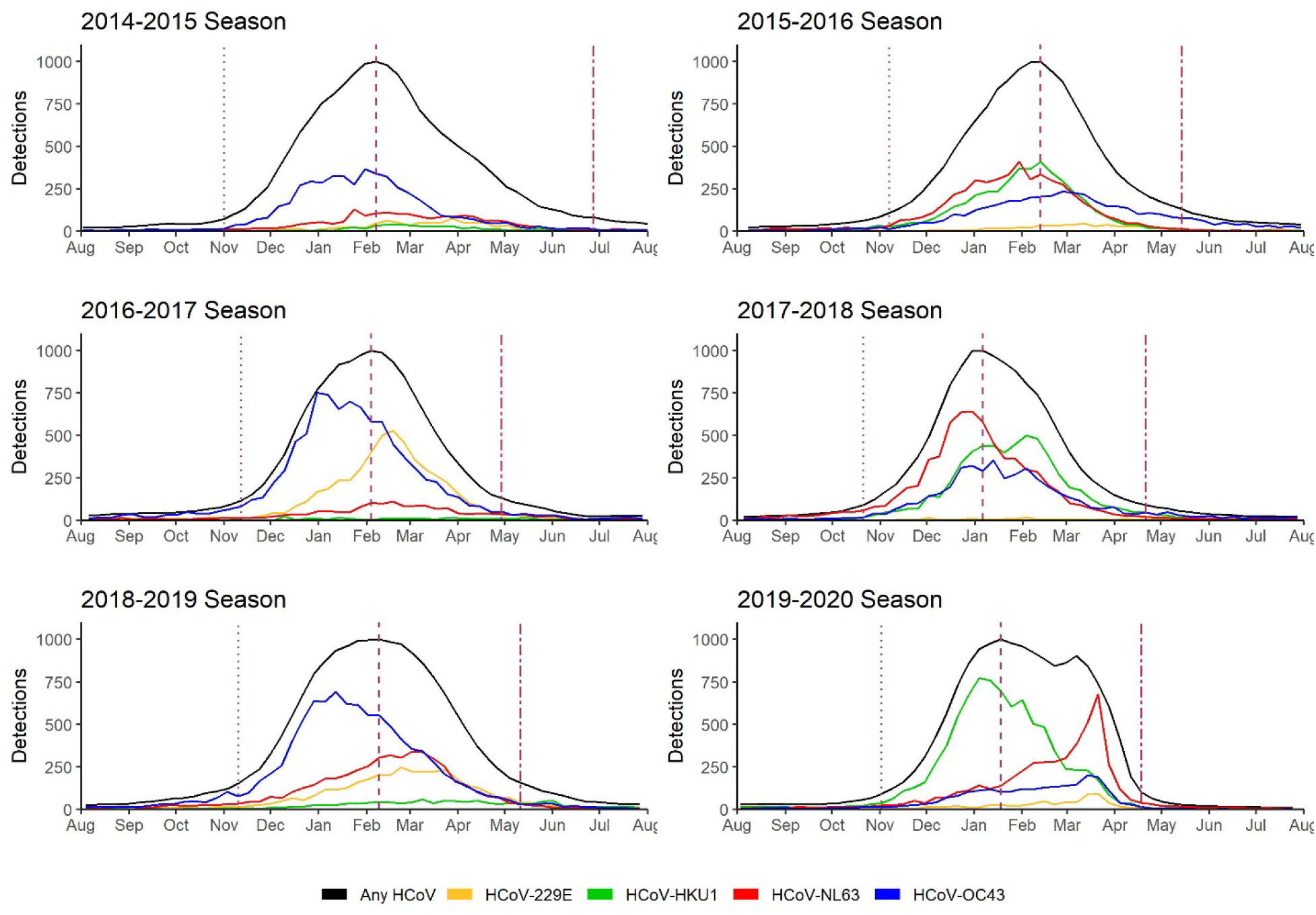
Total detections of the four common human coronaviruses (HCoVs) including HCoV-229E, HCoV-HKU1, HCoV-NL63, and HCoV-OC43 by week and season from weekly aggregated data submitted to the National Respiratory and Enteric Virus Surveillance System (NREVSS), July 2014-July 2020. The three vertical dotted lines in each panel indicate the week of season onset, peak, and offset for all types combined (black line). These seasonal inflections were defined by the Retrospective Slope 10 method which uses a centered 5-week moving average of weekly detections with normalization to peak. The type-specific curves depict the actual number of detections while the black curve depicts specimens with any HCoV detections normalized to a peak of 1000.

**Figure 3:**
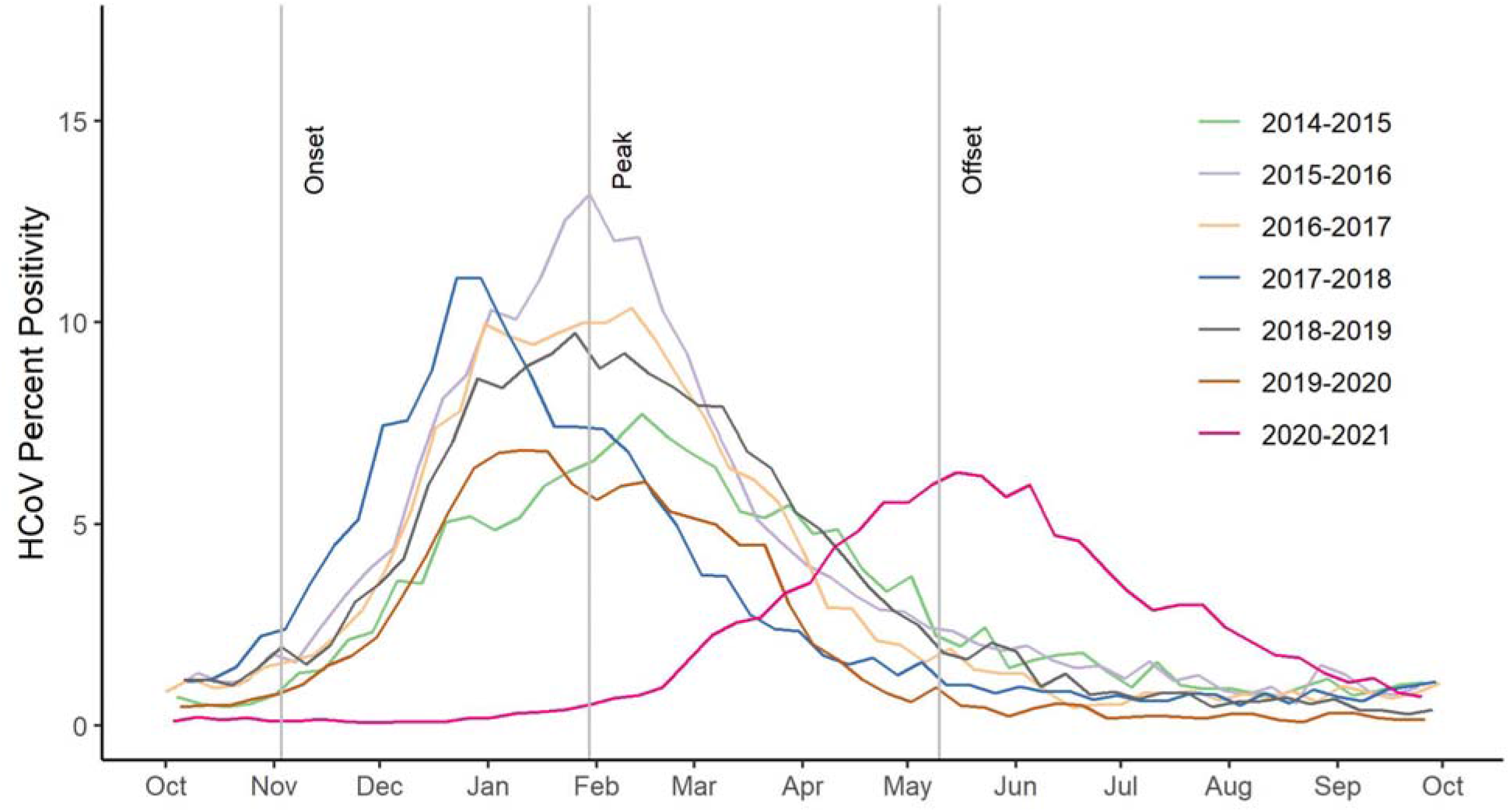
Percent positivity and seasonal characteristics of common human coronaviruses (HCoVs) by season in the United States from October 2014-September 2021 from weekly aggregated data submitted to the National Respiratory and Enteric Virus Surveillance System (NREVSS). The mean starting week dates for season onset, peak, and offset for all seasons except 2020-2021 are indicated by gray vertical lines based on the retrospective slope 10 method which uses a centered 5-week moving average of weekly detections with normalization to peak to define seasonal inflections. The average onset week for the six seasons spanning 2014-2020 is MMWR week 44, average peak week is MMWR week 04, and the average offset week is MMWR week 19. For the 2020-2021 season, the onset week is January 23 (MMWR week 03) and the peak week is May 22 (MMWR week 20) (not shown in figure).

**Table 1:** Onset, peak, and offset dates for the common human coronaviruses (HCoVs) including HCoV-229E, HCoV-HKU1, HCoV-NL63, and HCoV-OC43 from weekly aggregated data submitted to the National Respiratory and Enteric Virus Surveillance System (NREVSS), July 2014-July 2021. The dates for season onset, peak, and offset for all seasons are based on the retrospective slope 10 method which uses a centered 5-week moving average of weekly detections with normalization to peak to define seasonal inflections. Seasons were defined as starting on MMWR week 31 (early August) of each year through MMWR week 30 of the following year.

| Season | Onset Date (MMWR week) | Peak Date (MMWR week) | Offset Date (MMWR week) |
| --- | --- | --- | --- |
| 2014-2015 | November 1, 2014 (44) | February 7, 2015 (05) | June 27, 2015 (25) |
| 2015-2016 | November 7, 2015 (44) | February 13, 2016 (06) | May 14, 2016 (19) |
| 2016-2017 | November 12, 2016 (45) | February 4, 2017 (05) | April 29, 2017 (17) |
| 2017-2018 | October 21, 2017 (42) | January 6, 2018 (01) | April 21, 2018 (16) |
| 2018-2019 | November 10, 2018 (45) | February 9, 2019 (06) | May 11, 2019 (19) |
| 2019-2020 | November 2, 2019 (44) | January 18, 2020 (03) | April 18, 2020 (16) |
| 2020-2021 | January 23, 2021 (03) | May 22, 2021 (20) |  |

Among the alpha HCoVs, HCoV-NL63 was the predominant type during four of the seven seasons (Figure 1C). When either HCoV-229E or HCoV-NL63 were above 4% positivity, the other alpha HCoV had <1% positivity (Figure 1C). Among the beta HCoVs, HCoV-OC43 was the predominant type in a biennial pattern alternating with HCoV-HKU1 (Figure 1D) except for 2017-2018 when they were co-dominant. When either beta HCoV peaked above 4% positivity, the other beta HCoV circulated at low levels (<1% positivity), except for the 2015-2016 season (Figure 1D). Across U.S. census regions, patterns in the predominant HCoV type were similar to national patterns (Figure 4).

**Figure 4:**
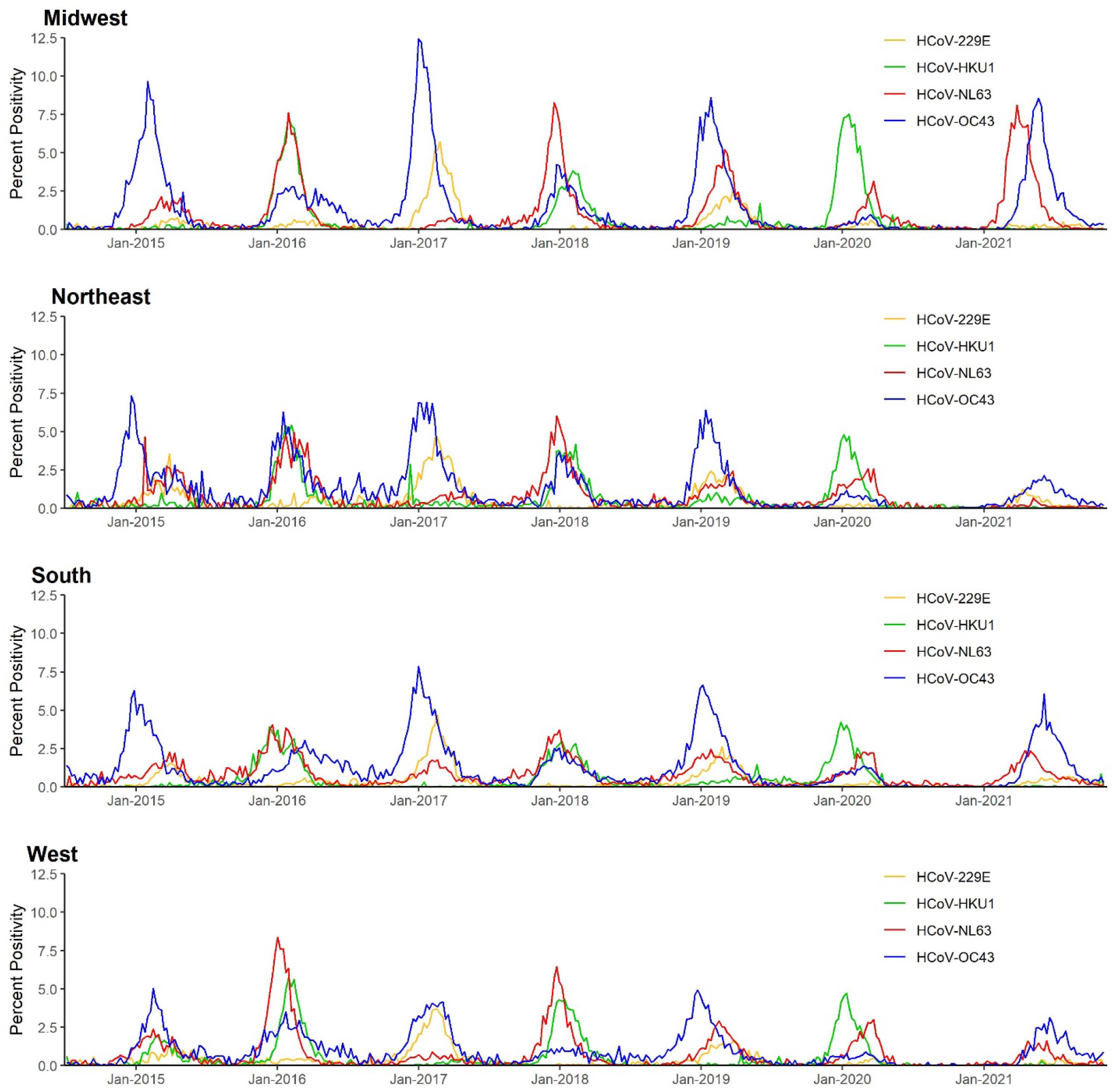
Percent positivity of the four common human coronaviruses (HCoVs) including HCoV-229E, HCoV-HKU1, HCoV-NL63, and HCoV-OC43 by week and United States Census region (Midwest, Northeast, South, and West) from weekly aggregated data submitted to the National Respiratory and Enteric Virus Surveillance System (NREVSS), July 2014-November 2021.

In 82,768 specimens from PHLIP tested for the four HCoVs between June 29, 2014 and November 29, 2021, 5,204 (6.3%) had any HCoV detection (Table 2). In 80,574 PHLIP specimens with information on sex the percentage of specimens with a HCoV detection was similar among males 6.0% (2617/43694) and 6.2% (2284/36880) females. Of the four types, HCoV-OC43 had the highest percentage of detections in children <1 year old and adults aged 66-100 (Table 2). HCoV-229E had the highest mean age and the lowest percentages in children 5 and under. Of specimens with a detected HCoV, 64/5204 (1.2%) had more than one HCoV type detected, and 1132/4685 (24.2%) had another respiratory virus detected, with influenza co-detections most common (9.8%) (Table 3).

**Table 2:**
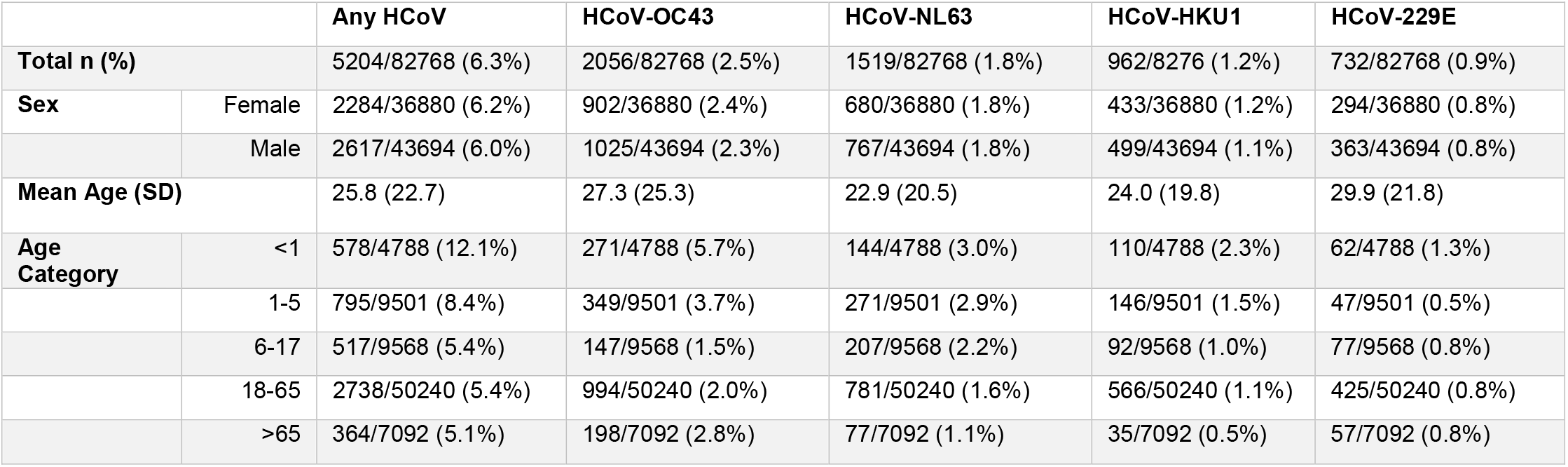
Percent positivity of the four common human coronaviruses (HCoVs) by sex and age categories, Public Health Laboratory Interoperability Project (PHLIP), July 2014-November 2021

|  |  | Any HCoV | HCoV-OC43 | HCoV-NL63 | HCoV-HKU1 | HCoV-229E |
| --- | --- | --- | --- | --- | --- | --- |
| **Total n (%)** |  | 5204/82768 (6.3%) | 2056/82768 (2.5%) | 1519/82768 (1.8%) | 962/8276 (1.2%) | 732/82768 (0.9%) |
| **Sex** | Female | 2284/36880 (6.2%) | 902/36880 (2.4%) | 680/36880 (1.8%) | 433/36880 (1.2%) | 294/36880 (0.8%) |
|  | Male | 2617/43694 (6.0%) | 1025/43694 (2.3%) | 767/43694 (1.8%) | 499/43694 (1.1%) | 363/43694 (0.8%) |
| **Mean Age (SD)** |  | 25.8 (22.7) | 27.3 (25.3) | 22.9 (20.5) | 24.0 (19.8) | 29.9 (21.8) |
| **Age Category** | <1 | 578/4788 (12.1%) | 271/4788 (5.7%) | 144/4788 (3.0%) | 110/4788 (2.3%) | 62/4788 (1.3%) |
|  | 1-5 | 795/9501 (8.4%) | 349/9501 (3.7%) | 271/9501 (2.9%) | 146/9501 (1.5%) | 47/9501 (0.5%) |
|  | 6-17 | 517/9568 (5.4%) | 147/9568 (1.5%) | 207/9568 (2.2%) | 92/9568 (1.0%) | 77/9568 (0.8%) |
|  | 18-65 | 2738/50240 (5.4%) | 994/50240 (2.0%) | 781/50240 (1.6%) | 566/50240 (1.1%) | 425/50240 (0.8%) |
|  | >65 | 364/7092 (5.1%) | 198/7092 (2.8%) | 77/7092 (1.1%) | 35/7092 (0.5%) | 57/7092 (0.8%) |

**Table 3:**
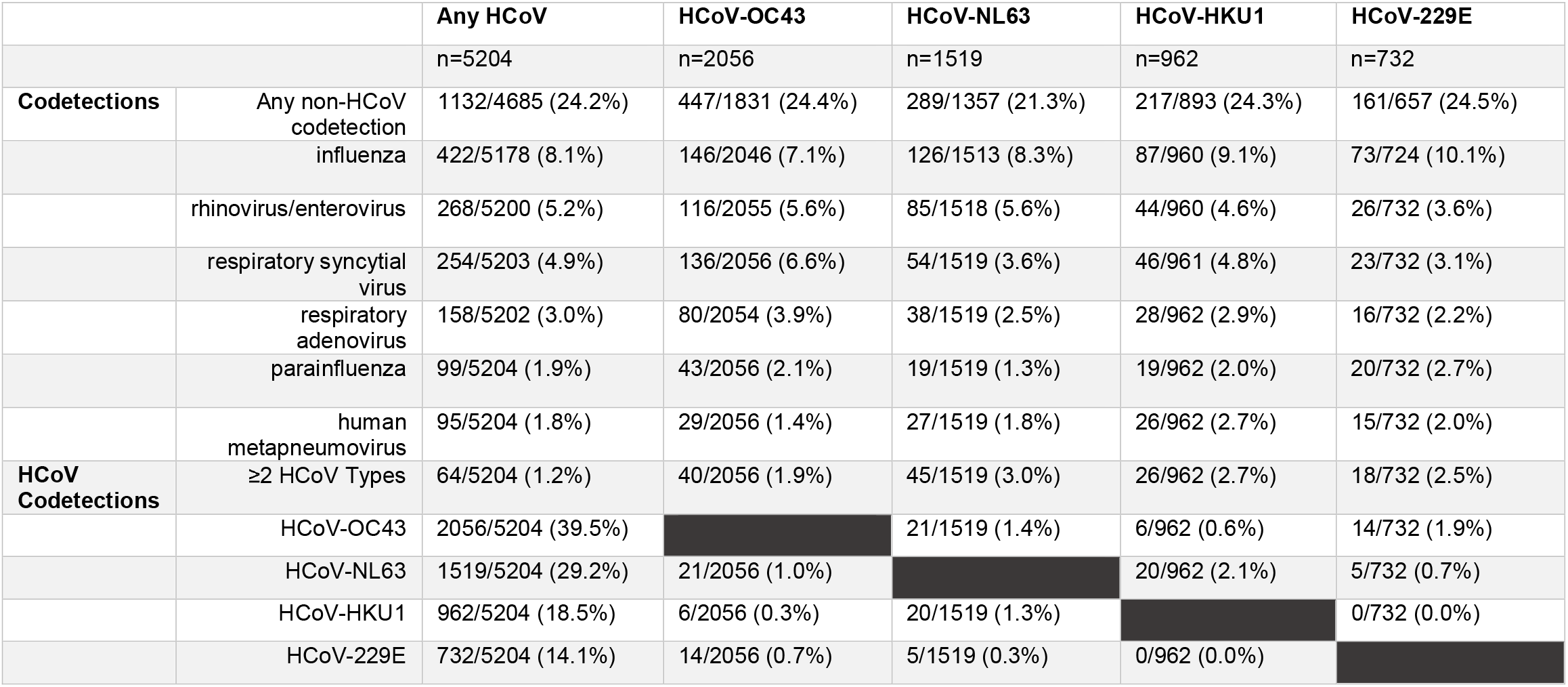
Respiratory codetections by common human coronavirus (HCoV) type, Public Health Laboratory Interoperability Project (PHLIP), July 2014-November 2021

## Discussion

During the last six seasons of HCoV circulation before the COVID-19 pandemic in 2020-2021, the relative consistency in timing of the seasonal onset, peak, and offset indicates expected patterns in the seasonality of HCoVs in the United States. The predominant type varied from season to season but at least one alpha HCoV and one beta HCoV circulated each season, often in a biennial pattern, as seen in other northern latitude countries (4). This biennial pattern may reflect cross-immunity and waning population-level immunity from prior infections to alpha and beta HCoVs (5), as serologic and human studies suggest immunity to reinfection lasting around one year (6, 7).

Cross-reactive binding and neutralizing antibodies seem to be higher among common HCoV types within a genera (8). SARS-CoV-2 cross-reactive serum antibodies were present in sera prior to the COVID-19 pandemic, likely attributable to cross-immunity from prior HCoV infections, but have not been shown to be protective against SARS-CoV-2 infection (9). Similarly, non-neutralizing antibodies to the common betacoronaviruses are boosted upon SARS-CoV-2 infection (9), but potential impact of SARS-CoV-2 immunity on seasonal HCoV circulation is yet unknown.

The seasonality of HCoVs is likely due to a combination of viral, host, and environmental factors. In temperate climates, HCoVs are observed to circulate during the winter months (10, 11); more variation and less predictable seasonality has been described in tropical regions compared to temperate regions (4, 12). Colder temperatures are thought to improve the stability of enveloped viruses. Additionally, lower temperatures lead to drying of airways, and can increase host susceptibility to infection. Environmental factors can also lead to behavior change which impacts the spread of HCoVs such as from more indoor human contact during winter months (13). Similarly, other widespread behavior changes could alter the seasonality of HCoV circulation.

The 2020-2021 HCoV season had a different pattern of circulation compared to prior seasons; the onset was delayed by over 2.5 months with an extended duration to peak. The 2020-2021 season offset could not be determined, as the number of detections had not fallen to low enough levels at the time of this analysis. In the United States, RSV and PIVs had delayed 2020-2021 seasonal starts while influenza and HMPV circulation was attenuated altogether (2, 14). RV/EV and HAdV activity was lower than usual at the beginning of season but activity increased to pre-pandemic levels later in the season. These changes are likely attributable in part to implementation of COVID-19 pandemic mitigation measures, such as decreased domestic and global travel, use of masks, school and office closures, and physical distancing (2).

Certain clinical and phenotypic differences from the four seasonal HCOVs have been previously observed, including differences in sex distribution, pathogenicity, and age. HCoV-OC43 has been reported as the most prevalent of the four common HCoVs consistent with this analysis (11). Male gender was associated with a higher odds of HCoV specimen positivity in a prior study (15), but males did not have a higher likelihood of having a HCoV detection in the current study.

HCoVs circulate seasonally with other respiratory viruses, including RSV, influenza, and rhinoviruses; coinfections are not uncommon (16, 17). We similarly show high levels of codetections (22%), particularly with influenza, which is likely an underestimate because only influenza detections from respiratory panels are included in the PHLIP dataset. Further work is needed to understand mechanisms of viral interference, and role of viral coinfections in the pathophysiology of illness and circulation of respiratory viruses.

There are several limitations to this investigation. Changes in testing patterns for respiratory viruses during the 2020-2021 season occurred due to delayed routine healthcare and an emphasis on SARS-CoV-2 testing, which impacts comparison to earlier seasons. The representativeness of codetections reported (i.e., true burden of illness) could not be evaluated because this PHLIP platform does not include reasons for testing (such as symptomatic disease); positive detections may be more likely to undergo reporting compared to negative detections. The NREVSS platform represents a geographically heterogenous subset of all U.S. laboratories but may not be nationally or locally representative. Furthermore, types of laboratory participation and the process for obtaining the subset of specimen level data for PHLIP are not fully comparable with the overall NREVSS platform.

Based on this analysis, a typical common HCoV season in the United States generally starts between October and November, peaks towards the end of January, and ends between April and June. This knowledge of expected seasonal variation in HCoV circulation is useful for public health preparedness and clinical management of patients. Clinicians and the public health community should be aware that patterns of HCoV circulation changed in 2020-2021, and that future seasons may also deviate from trends observed before the COVID-19 pandemic.

## Data Availability

The NREVSS dataset use in this seasonality analysis is included with this submission.

## Acknowledgements

We thank Phillip Salvatore for his assistance.

## Biographical Sketch

Melisa M. Shah is an Epidemic Intelligence Service Officer in the Respiratory Viruses Branch at the Centers for Disease Control and Prevention and an adult infectious diseases physician. Her research interests include the circulation of viral diseases and the natural history of SARS-CoV-2.

